# Biobank-scale Bayesian TWAS reveals splicing-mediated mechanisms of complex disease

**DOI:** 10.1101/2025.09.19.25336165

**Authors:** Alex Tokolyi, Saikat Banerjee, David A Knowles

## Abstract

Alternative splicing is a key mechanism by which genetic variation contributes to human phenotypic diversity and disease risk, yet its incorporation into large-scale genetic studies remains limited. Here we utilize blood RNA-seq from 4,732 European-ancestry individuals in the INTERVAL cohort to construct genetic scores for junction-based splicing phenotypes, 13,851 of which showed R^2^ > 0.01 in withheld individuals. These models were used to predict splicing in the UK Biobank White-British subset (UKBB; n=408,590) and All of Us individuals of diverse ancestries (AoU; n=201,958), enabling splicing transcriptome-wide association studies (sTWAS) across 1,129 harmonized disease phenotypes. We identified 4,966 splicing-disease associations, of which 67.3% were shared across cohorts. To account for confounding by co-regulation and linkage disequilibrium, we adapted a Bayesian joint fine-mapping framework to prioritize 1,277 likely causal splicing-disease associations across 494 genes, revealing mechanisms underlying immune, vascular, and metabolic traits. Fine-mapped associations included exon-skipping events in *GSDMB* and splicing in untranslated regions in *STAT6* and *IL18RAP* for asthma, unannotated splicing events in *PIEZO1* and *PPP3R1* for vascular traits, and alternative exons in *BAK1* and *TNFRSF14* for celiac disease. Our study demonstrates that common genetic variation influencing splicing substantially shapes complex trait architecture and provides a scalable, statistically rigorous framework to uncover transcriptional disease mechanisms.

## Main

Alternative splicing is a fundamental mechanism of eukaryotic gene regulation, contributing to transcriptomic and proteomic diversity by allowing a single gene to produce multiple RNA isoforms, as well as regulating gene dosage. While alternative splicing is pervasive across the human genome, with over 95% of multi-exon genes exhibiting evidence of splicing variation^1^, the genetic architecture underlying this process is complex and highly context-dependent^2^. Splicing quantitative trait loci (sQTLs) capture common regulatory variants that influence splice junction usage, and are increasingly recognized for their contribution to disease risk, particularly when they modulate protein-coding potential, regulatory element inclusion, or dosage^3–5^.

Large-scale efforts have catalogued sQTLs across tissues, with studies such as GTEx^2^ and INTERVAL^4^ identifying thousands of splicing events influenced by local (*cis-*) and distal (*trans-*) genetic variants. These insights have laid the groundwork for transcriptome-wide association studies (TWAS), which integrate genetic and transcriptomic data to assess whether the genetically regulated component of gene expression or splicing is associated with complex traits^6,7^. While expression-based TWAS have illuminated thousands of disease-associated loci^8–10^, splicing has only recently begun to receive comparable attention. This gap is particularly notable given growing evidence that splicing variation is enriched for disease-associated loci and often provides independent or complementary signals to gene expression^3,11^.

The incorporation of splicing into genetic prediction models and biobank-scale analyses presents both computational and methodological challenges. Splice junction phenotypes are higher-dimensional, more sparse, variable across individuals and tissues, and differ in their quantification approaches^3^. Additionally, predictive modeling of splicing phenotypes requires accounting for a genetic architecture which is less well studied than that of total gene expression. Despite these complexities, splicing is likely an important mediator of genetic risk, particularly for the discovery of causal genes that are missed by expression-only analyses (false negatives) or attributed to incorrect transcriptional mechanisms (false positives).

Efforts to integrate splicing into genetic risk models have been limited in scale, particularly across diverse populations and biobanks. Given the scale and heterogeneity of human disease phenotypes, biobank resources such as the UK Biobank^12^ (UKBB) and All of Us^13^ (AoU) offer an unprecedented opportunity to explore the role of genetically regulated splicing in health outcomes across populations. To enable such analyses, robust splicing genetic scores are needed that are accurate, generalize across cohorts, and are sufficiently scalable to handle tens of thousands of traits in hundreds of thousands of individuals.

Here, we develop and benchmark splicing genetic scores across 111,937 splicing phenotypes (both known and *de novo* detected) derived from whole blood RNA-seq in the INTERVAL^4^ cohort (n=4,732 individuals). Using Bayesian ridge regression we generate polygenic scores for 13,851 splicing events demonstrating withheld predictive accuracy (R^2^ > 0.01), and impute these into both the UKBB and AoU cohorts. Leveraging harmonized PheCodes^14,15^ across biobanks, we perform a splicing TWAS on 1,129 disease phenotypes using case-control logistic regression, identifying thousands of splicing-disease associations and highlighting shared signals across genetically distinct populations. We then adapt a Bayesian joint fine-mapping framework^16^ to resolve pleiotropic effects, integrating both our splicing and external gene expression scores^10^ to prioritize likely causal splicing events **(Figure 1)**. Our open access results demonstrate that common variation modulating splicing contributes broadly to disease risk, revealing new insights into the functional basis of complex traits (http://stwas.nygenome.org).

**Figure 1.**
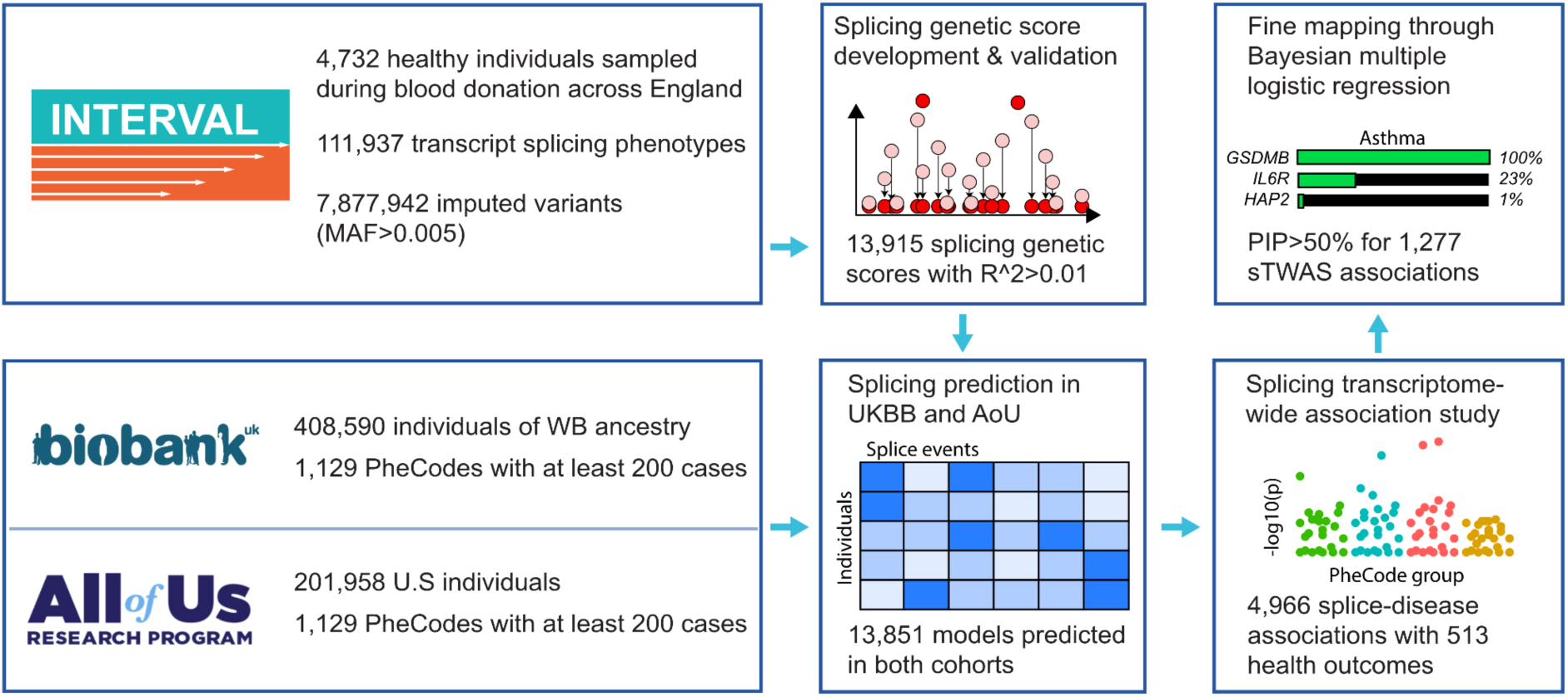
Overview of the cohorts and data used for this study, along with the main analytical approaches.

## Results

### Creation of splicing genetic scores and biobank imputation

We obtained 111,937 splicing phenotypes and associated *cis-* and *trans-*sQTLs assayed from 4,732 European-ancestry individuals and associated imputed genotypes from the INTERVAL RNA-seq analysis^4^. We retained splicing events that were wholly contained within a gene body, were outside the extended HLA region^17^, and had at least one significant sQTL for a non-ambiguous SNP. We utilized Bayesian ridge regression to train splicing genetic scores (**Methods**) as it has been shown to be robust for transcriptomic traits, as well as computationally scalable to the large number of traits and individuals selected here^10^. We observed large variation in predictive accuracy (R^2^), likely reflective of varying heritability, across splicing genetic scores (**Figure 2A, Table S1**), with 14,783 splicing phenotypes with mean internal R^2^>0.01, 3,797 with R^2^>0.1, 1,118 with R^2^>0.3, and 422 with R^2^>0.5 (**Figure 2B**). Additionally, the inclusion of *trans*-sQTLs led to an additional 322 splicing events with an internal R^2^>0.01 **(Figure 2C)**, and an increase in R^2^ for 739 others. Using a withheld 20% subset of the INTERVAL cohort, and the combined *cis* and *trans* models, 13,851 splicing genetic scores (in 4,137 unique genes) had a mean withheld test-set R^2^>0.01 and passed our inclusion criteria (**Methods**): we chose these to predict in the UK Biobank and All of Us (**Figure 2D**). Imputed splicing events were predominantly within protein coding genes and mostly previously annotated, though 30.6% had a novel donor or acceptor site (**Figure 2E, Table S2**). Just over a third (35.4%) excised a protein coding portion of the transcript, leading to a higher likelihood of downstream proteomic consequences. Disease phenotypes were mapped to PheCodes^15^ (**Methods**), selecting for those with a minimum of 200 cases (n=1,129 PheCodes) in both the UK Biobank and All of Us cohorts (**Figure 2F, Table S3)**. Due to differences in health systems and EHR records, there was a large variation in case/control ratio, with that of the All of Us cohort being a median 5.48x higher than that of the UK Biobank **(Figure S1)**, consistent with previous observations^18^.

**Figure 2.**
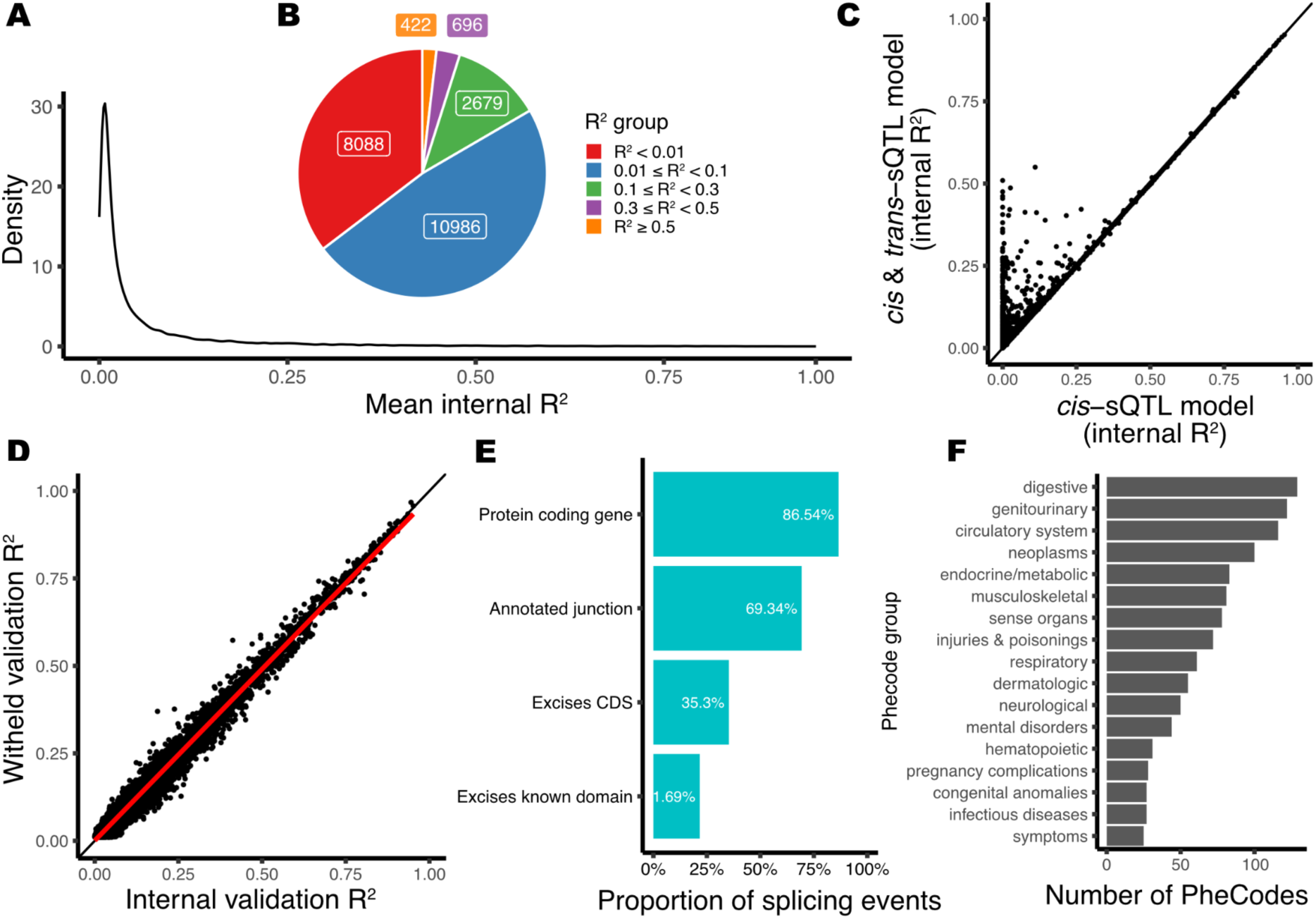
Validation of genetically predicted splicing and selected PheCodes for sTWAS analysis. **A)** Density plot showing the mean internal R^2^ for all tested splicing genetic scores, with an inset **B)** pie chart showing the quantity of splicing scores at various ranges of R^2^. **C)** The relationship between internal R^2^ scores between a *cis*-sQTL only model (x) and a model which additionally includes *trans-*sQTLs (y). **D)** Alignment of internal validation R^2^ (x) with that computed on the withheld test subset of the INTERVAL cohort (y), black line equates to y=x, red line is the slope of y∼x. **E)** Overlap of the selected splicing events with main transcriptomic and proteomic features (overlap in blue, percent inset in white). **F)** Quantity of filtered PheCodes by group classification.

### Splicing TWAS reveals splicing-disease associations

Using our 13,851 splicing genetic scores, we performed a TWAS on 1,129 PheCodes in both the UK Biobank and All of Us cohorts using a case/control logistic regression (**Methods**). At an FDR of 0.05 we detected 2,479 significant associations between 1141 splicing events and 264 PheCodes in the UK Biobank (n=408,590), and 573 significant associations between 325 predicted splicing events and 145 PheCodes in All of Us (n=201,958), likely owing to the reduced sample size. To assess the sharing of sTWAS results between cohorts we performed multivariate adaptive shrinkage (mash) between the summary statistics of both cohorts (**Methods**), revealing that 67.3% of associations between the two cohorts were shared (within a factor of 0.5) (**Figure 3B, Table S4)**, with the remainder predominantly possessing a larger effect in the UKBB. This level of sharing is remarkably high, given that there were both differences in sampled populations and measurement/diagnosis between the two biobanks. Given the high levels of sharing, we performed a fixed-effects meta-analysis on the sTWAS results from both cohorts, yielding 4,966 significant associations (FDR<0.05), between 2,107 splicing events (in 1,012 genes), with 513 PheCodes **(Figure 3A, Table S5)**. The fixed-effects meta-analysis of UK Biobank and All of Us TWASs showed well-calibrated test statistics (λ_meta=1.097; **Figure S2**), consistent with the modest inflation expected for highly polygenic traits. While we chose to limit splicing events to those with a withheld R^2^ threshold of >0.01 to maximize discovery, we conducted a sensitivity analysis of more stringent R^2^ thresholds (**Figure S3).** This showed that the *proportion* of significant sTWAS associations increased at more stringent thresholds, demonstrating that higher-quality models were more likely to yield a significant association.

**Figure 3.**
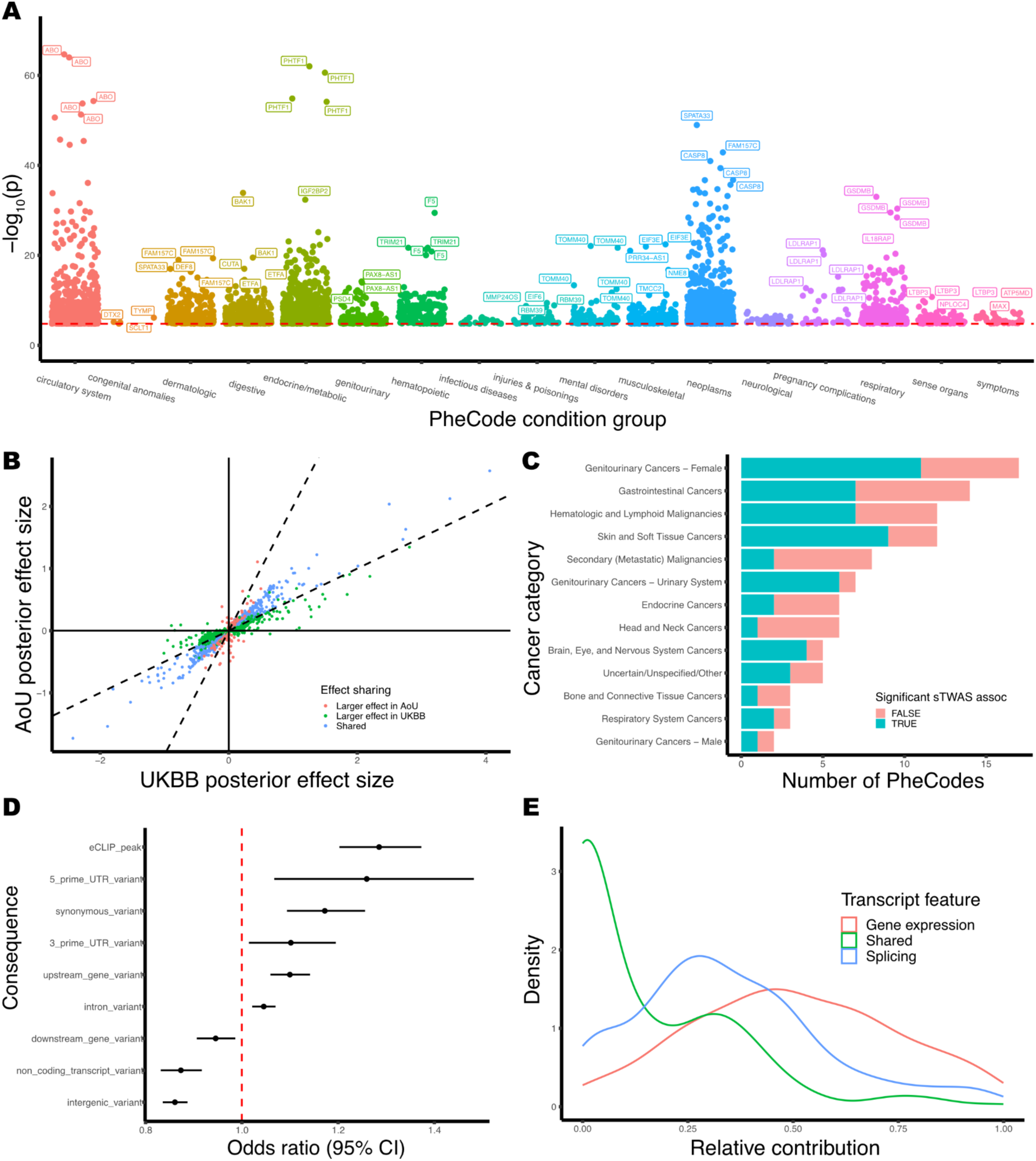
Summary of splicing TWAS results between UKBB and AoU. **A)** Manhattan-style plot of the sTWAS meta-analysis between UKBB and AoU, grouped and coloured by PheCode group (x) and significance (y), with the gene names of the 5 most significant splicing events per group labelled. **B)** mash analysis of the effect sharing of sTWAS results, showing the UKBB (x) and AoU (y) sTWAS posterior effect sizes. These are coloured by the unique or shared effect status between the two cohorts. **C)** Quantity of cancer PheCode groups which have and do not have significant sTWAS association. **D)** Enrichment of SNP consequences for SNPs of splicing event models with a significant sTWAS association. **E)** The relative independent and shared contribution (green) of significant splicing events (blue) and gene expression (red) to tested PheCodes, with clamping (negative fractions set to zero and sum renormalized to one).

Neoplasms made up a large proportion of the significant sTWAS results, so we assessed the number of associations in different broad cancer types (**Figure 3C**). We observed that while female genitourinary cancers were the most commonly tested neoplasm group, those relating to skin and soft tissue cancers were the most likely out of the cancer PheCodes to have a significant association (91.7%). We compared our sTWAS to the current largest breast cancer sTWAS^19^ which included breast cancer RNA-seq samples from 652 individuals and a case-control cohort of 426,834 individuals (n=178,534 cases). Our sTWAS revealed associations with breast cancer for splicing in 23 genes, 5 of which (*KANSL1, LSP1, CASP8, FDPS, SCAMP3*) were also observed in their analyses (p=2.21e-06), and 19 of which were collated by the authors as previous expression TWAS associations for breast cancer (p=2.68e-25).

### Global mechanisms of splicing effects on disease

To determine potential global mechanisms for the genetic variants underlying disease risk through splicing, we analysed SNPs present in the models of at least one significant splicing-disease association with ENCODE RNA binding protein (RBP) data^20^. SNPs that contributed to splicing phenotypes that were significant in our TWAS were 26% more likely to be contained within an RBP peak (p=1.03e-8, OR=1.26, 95% CI: 1.17-1.37, Fisher’s exact test). We additionally assessed VEP variant consequences for these SNPs, observing 8 other significantly enriched variant consequences (FDR<0.05, **Figure 3D**). We then sought to analyse the characteristics of splicing events that had a significant sTWAS association, using the 13,851 tested events as a background set. These were annotated using the Leafidex pipeline **(Methods)**, which detects overlaps with transcriptomic features from GENCODE^21^, and proteomic features from UniProt^22^ and Pfam^23^ (**Table S2)**. Of the 15 characteristics tested, 4 were significant (FDR<0.05), including splicing events that excise extracellular domains, known domains (Pfam), and those that overlap exonic or CDS regions (**Figure S4**).

### Splicing provides added information to expression-based TWAS

We imputed 13,668 gene expression genetic scores developed by Xu *et. al.*^10^ into the UK Biobank and All of Us cohorts. In a single cohort for comparison (UKBB), an expression TWAS (eTWAS) was performed using the same model and covariates using the same methodology as for the sTWAS, to assess the added contribution of transcript splicing to the eTWAS gene-PheCode associations. We observed 3,502 significant gene-PheCode pairs, compared to 2,600 in the sTWAS. Interestingly, only 194 of these pairs overlap, suggesting that the majority of the sTWAS results are capturing gene-disease relationships that are unique to, and independent of gene expression. To assess the relative contribution of splicing and gene expression to disease risk, we jointly modeled significant traits from the expression TWAS and splicing TWAS for each disease within UKBB. Across 204 PheCodes with both significant expression and splicing associations, we decomposed the unique and shared effects of each modality (**Figure 3E, Table S6)**. On average, gene expression explained 48.9% of the transcriptomic contribution, splicing accounted for 31.7%, and a shared component contributed 5.4%, though proportions varied widely across traits **(Figure S5)**. These results indicate that splicing contributes substantially and independently to disease architecture, complementing gene expression and highlighting the value of modelling both sources of transcriptomic variation.

### Transcriptome fine-mapping of splicing TWAS results with Bayesian joint analysis

The basic TWAS approach above, where we test each splicing event-trait pair separately, is known to be confounded by co-regulation/pleiotropy and linkage disequilibrium^24,25^. As the sTWAS used a univariate logistic regression model testing only a single predicted splicing event and PheCode, these were regularly (308/513 PheCodes) associated with multiple predicted splicing events within the same linkage disequilibrium (LD) block. Total gene expression is also tightly linked with splicing, due to the specific upregulation of individual transcript isoforms or splicing-induced nonsense mediated decay. Thus splicing and expression phenotypes for a gene may be co-regulated/show pleiotropy. To untangle these effects and prioritise individual splicing events, we adapted a Bayesian joint logistic regression model previously developed for GWAS fine-mapping (B-LORE^16^) to TWAS traits (**Methods**). To allow for prioritization between splicing and total gene expression, including that of nearby (and potentially *cis-*associated) genes, we imputed 13,668 gene expression genetic scores developed by Xu *et. al.*^10^ into the UK Biobank and All of Us cohorts using the same methodology as for splicing. We emphasize that the covariates in the B-LORE model are splicing and gene expression events, rather than SNPs as in typical “fine-mapping”, and the resulting posterior inclusion probabilities (PIP) represent the evidence for a causal link from splicing event to trait. For each significant splicing-PheCode association, we aggregated all predicted splicing and gene expression scores located within the same LD block (**Methods**) and fit a Bayesian joint logistic regression model for each significant PheCode. A Bayesian meta-analysis was then performed across the two cohorts to compute a meta PIP per splicing-PheCode pair. In total, we observed 1,277 pairs with PIP>0.5 for 779 unique splicing events (in 494 genes) and 191 disease PheCodes (**Table S5**). The PIPs exhibited a bimodal distribution, with most values clustering near 0 or 1, indicating that the model confidently distinguished between non-causal and likely causal features **(Figure 4A)**.

**Figure 4.**
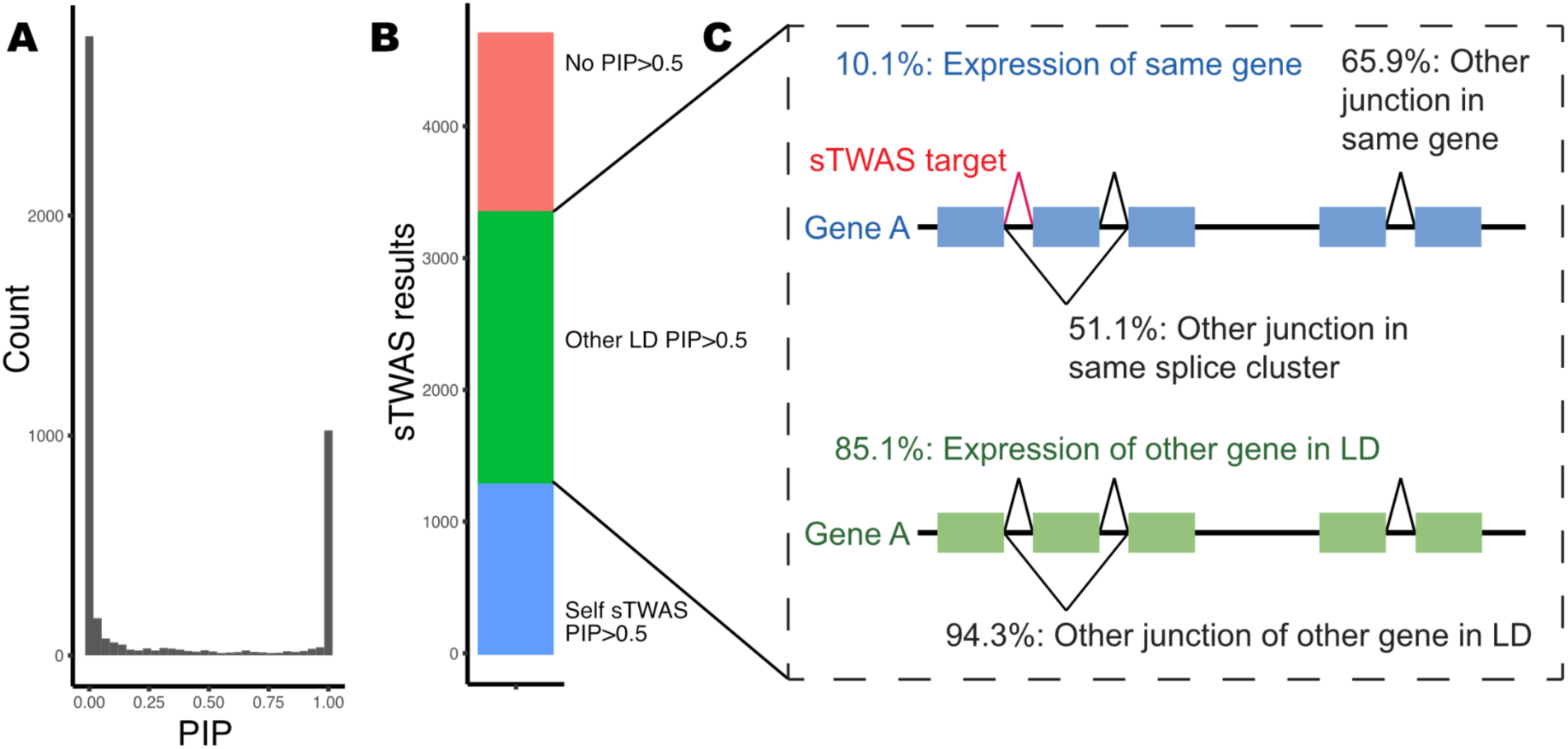
Bayesian fine-mapping of splicing TWAS results reveal causal transcriptomic sources. **A)** Distribution of PIP scores for significant sTWAS splicing events. **B)** Amount of sTWAS results with a causal PIP for the tested splicing event (“self PIP”, blue), for another transcriptomic source in the LD block (“other PIP”, green), or no detected causal source (“no PIP”, red). **C)** Schematic of other causal transcriptomic sources within the LD block of fine-mapped sTWAS results, either within the same gene (blue) or another gene in the LD block (green).

To explore the cross-ancestry portability of our sTWAS results, we quantified attenuation of TWAS effect sizes within All of Us by regressing Admixed American-ancestry and African-ancestry sTWAS betas on European-ancestry sTWAS betas (inverse-variance weighted). Among our high-confidence associations that were significant in meta-analysis and had fine-mapping support (PIP > 0.5), we observed mild attenuation (λ_AMR=0.90, λ_AFR = 0.86), indicating largely preserved effect sizes in non-European ancestries. In contrast, when considering all AoU European-ancestry significant associations, attenuation was more pronounced (λ_AMR = 0.55, λ_AMR = 0.56), consistent with reduced portability and greater heterogeneity among weaker or less well-resolved signals.

To investigate the causal source of the detected sTWAS signals when the associated splicing event was non-causal, we collected the gene expression and splicing events within the LD block that were allocated causal status (PIP>0.5). This was stratified into two gene expression categories (expression of the spliced gene, or another gene in the LD block), and 3 splicing categories (splicing event in same cluster, same gene, or in another gene in the LD block). Of the 4,700 events for which B-LORE converged, 1,277 (27.2%) had a PIP>0.5 for the original significant sTWAS splicing event, 1,355 (28.8%) had no detected causal source in the LD block, and for 2068 (44.0%) B-LORE suggests a different transcriptomic event as the causal mechanism **(Figure 4B)**. Of the latter category, the estimated cause was most commonly a different splicing event within another gene in LD (94.3%) and interestingly least often to be expression of the containing gene (10.1%, **Figure 4C**).

### Splicing mechanisms underlying immune, vascular and metabolic traits

Fine-mapped splicing-disease associations revealed mechanisms across immune-mediated, vascular, and metabolic traits among others, consistent with the broad relevance of the blood origin of our transcriptome data. For asthma, we identified 30 splicing events across 18 genes with meta PIP>0.5. An exon 5 skipping event in *GSDMB*, which abolishes the pyroptosis-inducing domain, was associated with reduced risk (p=9.7e^-34^, OR=0.81, PIP=1). While the 17q21 asthma locus has multiple candidate genes; our results provide evidence that *GSDMB* splicing variation, rather than *ORMDL3* or *GSDMB* gene expression, underlies the association **(Figure 5A)**, consistent with prior functional evidence^26^. A rare, partial loss-of-function variant in the Th2 transcription factor *STAT6* has been observed to be protective in asthma^27^. We observe opposing effects for splicing at the untranslated regions of *STAT6* (**Figure 5B**): excision of the 5′ UTR reduced risk (p=4.6e^-18^, OR=0.81, PIP=1), whereas a 3′ UTR event increased risk (p=1.44e^-12^, OR=1.26, PIP=1), in the absence of a gene expression signal. This may act through reducing STAT6 translation, and provides a mechanism for the impact of common variation acting through splicing within this gene. The *IL18RAP* locus (also containing the interleukin receptors *IL1RL1, IL1R1,* and *IL18R1*) has been previously implicated in asthma without a straightforward mechanistic explanation^28^. We identified a 5′ UTR splicing event that decreased risk (p=3.0e^-24^, OR=0.86, PIP=1), providing a putative mechanistic interpretation of the GWAS signal. We additionally observed causal signals for total gene expression in 3 nearby interleukin genes (*IL1RL1*, *IL1R1* and *IL18R1*), though not expression of *IL18RAP*, suggesting a mechanism that interacts through multiple causal signals in both splicing and total expression at this locus **(Figure 5C)**.

**Figure 5.**
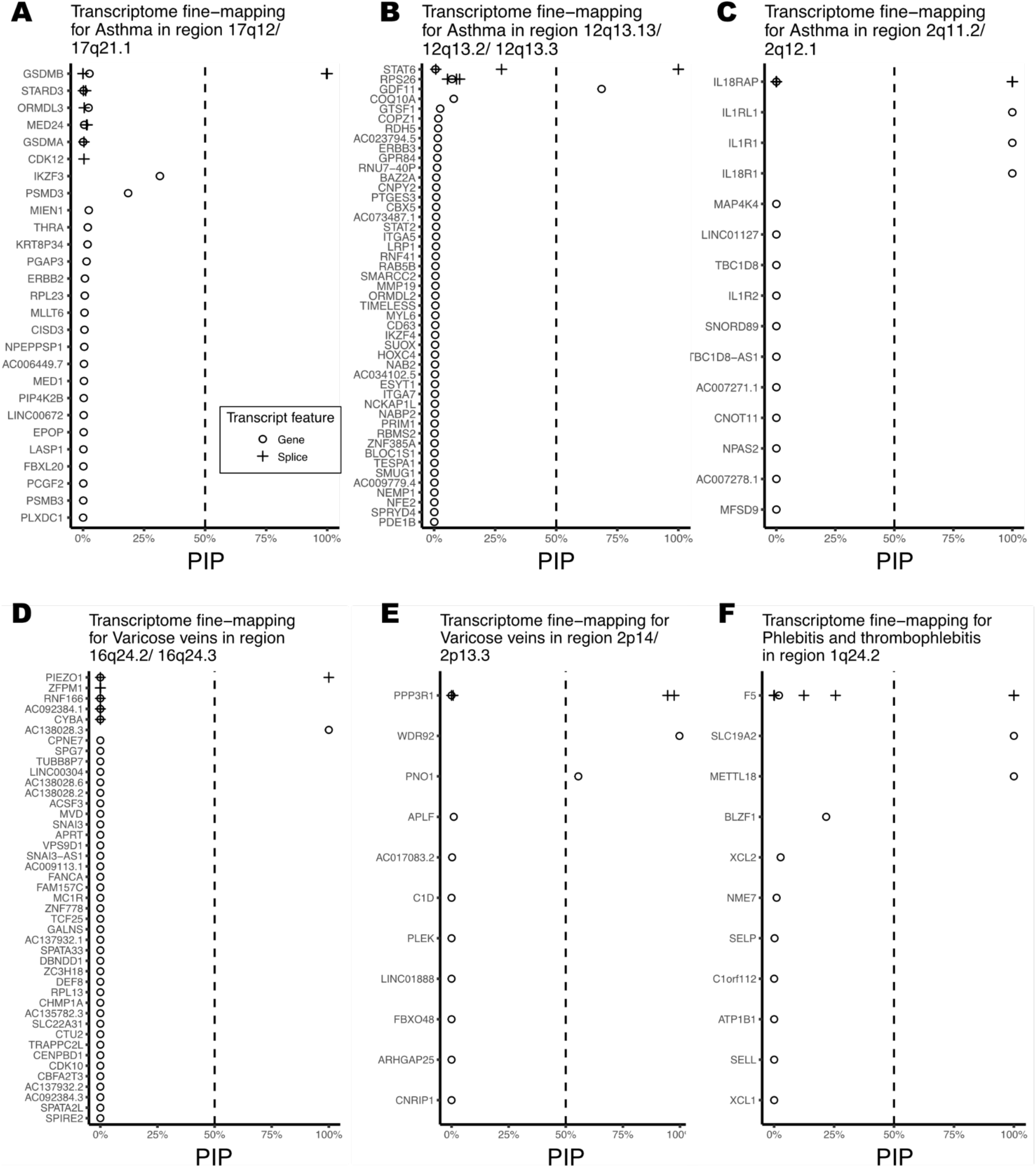
Transcriptome-wide fine-mapping of TWAS results. **A)** Tower plot showing the sTWAS fine-mapped PIPs for transcriptomic events (splicing events as “+”, gene expression as “◯”) for the LD block containing *GSDMB* with asthma risk. Tower plots as previously for **B)** the LD block containing *STAT6* with asthma risk, **C)** the LD block containing *IL18RAP* with asthma risk, **D)** the LD block containing *PIEZO1* with risk of varicose veins, **E)** the LD block containing *PPP3R1* with risk of varicose veins, and **F)** the LD block containing *F5* with risk of phlebitis and thrombophlebitis.

Among vascular traits we observed 346 sTWAS associations with fine-mapping evidence, the most of any PheCode group. For varicose veins, an unannotated splicing event in the 3′ UTR of *PIEZO1* decreased risk (meta P=3.5e^-22^, OR=0.53, PIP=1, **Figure 5D**), complementing rare coding variant evidence at this gene^29^. *PEIZO1* has known roles in erythrocyte volume regulation and thrombosis, so alternative isoforms containing this splicing event could be acting through altered blood cell or endothelial function. At *PPP3R1* which has prior genetic evidence for association with varicose veins^30^, two partially-annotated splicing events with opposing directions of effect were nominated as causal, with higher predicted expression of an annotated intronic splicing event decreasing risk (p=5.86e^-28^, OR=0.77, PIP=0.97, **Figure 5E**), while the other event, with a shared donor and novel acceptor, increased risk (p=9.56e^-28^, OR=1.26, PIP=0.95). As *PPP3R1* encodes a regulatory calcineurin complex subunit, these two splicing events could represent a delicate balance in calcineurin in this condition. Transcriptome-wide fine-mapping at this locus solely nominated the splicing of *PPP3R1*, along with the expression of two nearby genes. For coagulation traits, we identified seven previously unannotated but in-frame splicing events within exon 13 of *F5*, which encodes the B domain. Each event excises a segment divisible by three nucleotides, consistent with the generation of alternative isoforms that may act analogously, though more subtly, to the well-characterized FV-short isoform, which removes a much larger portion of exon 13 and is present at low but variable concentrations in most individuals^31,32^. These exon 13 events were predicted to be causal for disorders including thrombophlebitis (min p=1.48e^-16^, OR=0.84, PIP=1, **Figure 5F)** and several clotting deficiencies.

For metabolic and immune traits, splicing highlighted additional mechanisms. In celiac disease, a novel splicing event truncating the final coding exon of *BAK1* increased risk (meta P=1.3e^-34^, OR=3.40, PIP=1, **Figure 6A**), consistent with the modulation of pro-apoptotic activity^33^. We additionally observe a splicing event in *TNFRSF14*, encoding the pro-apoptosis receptor protein HVEM^34^, with known roles in intestinal inflammation^35–37^. This splicing event possesses with an annotated donor and novel acceptor excising part of coding exon 7, containing the cytoplasmic domain, and was observed to decrease the risk of celiac disease (p=2.2e^-6^, OR=0.71, PIP=1, **Figure 6B**). As this splicing event excises a coding portion of the receptor protein, it is possible that this splicing event modulates celiac disease through the signalling of this pro-apoptotic receptor. Hypothyroidism showed a large signal in our sTWAS, with fine-mapped association for 53 splicing events across 36 genes. Two alternative 5′ UTR splicing events in *PHTF1* displayed opposite effects on hypothyroidism risk (p=2.5e^-61^ & 7.3e^-55^; OR=1.74 & 0.53; PIP=1 & 1, **Figure 6C**), potentially acting through altered translational regulation. Together, these examples illustrate how common splicing variation refines the mechanisms underlying GWAS loci across diverse trait classes, with full associations detailed in **Supplementary Table S5**, and online in our portal (http://stwas.nygenome.org/).

**Figure 6.**
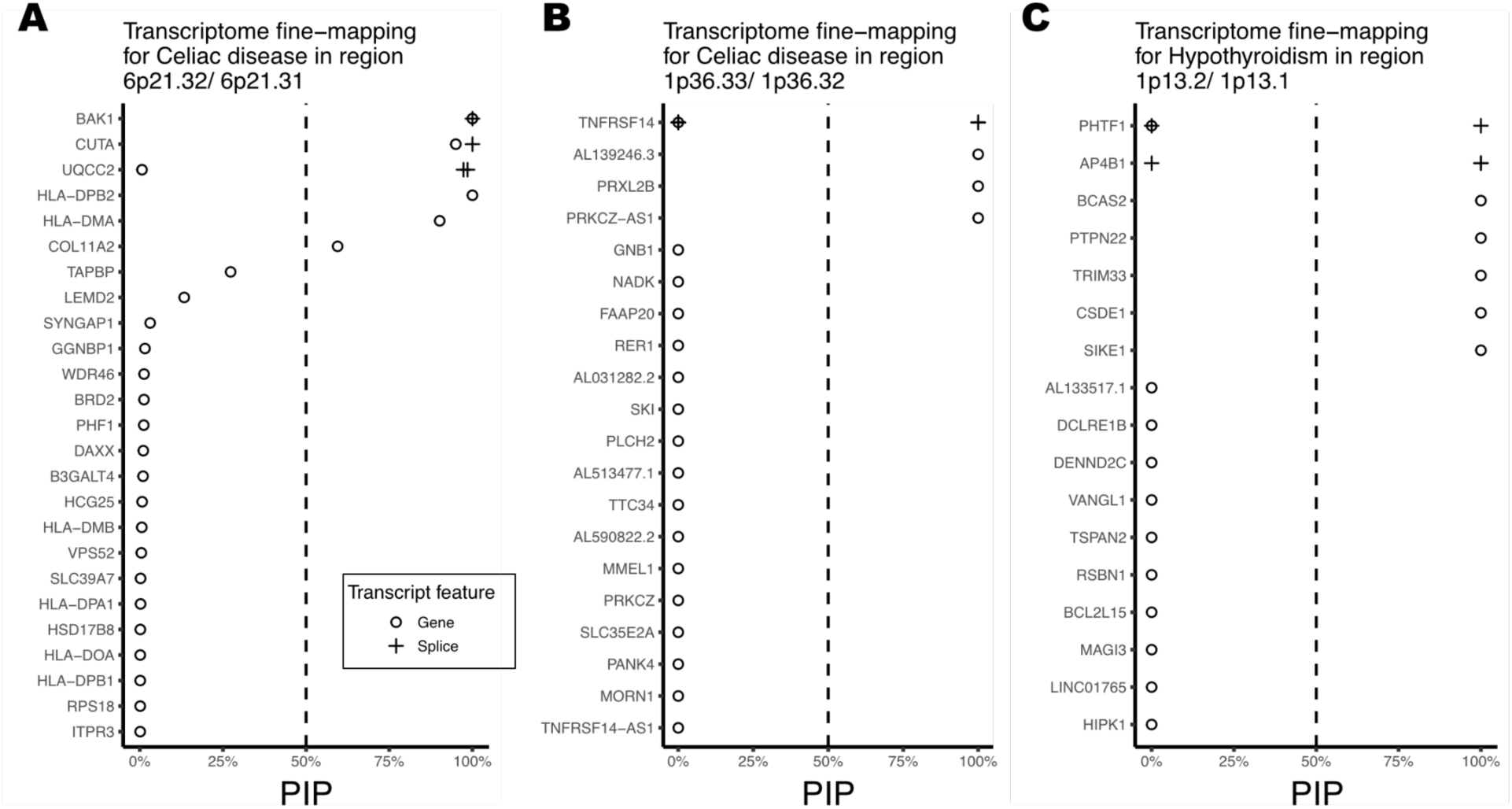
Transcriptome-wide fine-mapping of TWAS results. **A)** Tower plot showing the sTWAS fine-mapped PIPs for transcriptomic events (splicing events as “+”, gene expression as “◯”) for the LD block containing *BAK1* with celiac disease risk. Tower plots as previously for **B)** the LD block containing *TNFRSF14* with celiac disease risk, and **C)** the LD block containing *PHTF1* with hypothyroidism.

## Discussion

Our study contributes to the growing evidence that splicing is a major and previously underappreciated component of the genetic architecture of complex disease. By developing splicing genetic scores, performing a large-scale splicing TWAS across both the UK Biobank and All of Us cohorts, and applying a novel Bayesian joint fine-mapping approach, we identified over a thousand splicing-disease associations with high posterior probability of causally impacting disease risk. Integrative modelling of splicing and expression further demonstrated that splicing independently accounts for on average nearly one third of the transcriptomic contribution to disease risk, independent of gene expression. These findings establish splicing as a complementary and substantial dimension of genetic regulation, extending beyond expression-based models, and refining mechanistic interpretation of GWAS loci. Using our splicing features derived from whole blood, we were able to recapitulate both existing findings, such as *GSDMB* exon 5 splicing in epithelial cells impacting asthma risk^26^, multiple genes in a recent breast-cancer sTWAS^19^, as well as many new examples where splicing may explain GWAS risk loci without established mechanisms. Additionally, we note the functional importance of splicing annotations in deciphering their risk, through the observation of enrichment of the excision of topological domains and 5’ UTR and 3’ UTR splicing in disease. Notably, the splicing events we identified are often modulatory rather than outright destructive, implicating protein localization or receptor shedding rather than complete loss of gene function. Therapeutically, targeting of these specific splice junctions may provide more context specific effects than full gene knockouts, increasing specificity and decreasing the risk of off-target effects.

Previous TWAS studies have focused almost exclusively on total gene expression^6,7^. We demonstrate that the addition of splicing to these analyses adds additional, unique information to that of gene expression, and links to the growing recognition of post-transcriptional regulation in disease. While our analyses provide many novel insights, they should be interpreted in the context of several limitations. While we included splicing features quantified *de novo* to allow for those that are less common, our features do not represent the totality of the transcriptome. For example, the methods used did not consider alternative transcription start and end sites, which may also act as a mechanism through which genetic variation can exert its effect. Additionally, although the meta-analysis of UKBB and AoU increased power, differences in cohort selection and healthcare systems^18^ may have obscured some effects. Our reliance on healthy whole blood to compute splicing genetic scores, while possessing many shared genetic effects with other tissues^38^, may miss splicing effects which are tissue-specific in distal tissues, or disease-specific splicing events. Due to the nature of the short-read sequencing from which our splicing features were derived, we lack the full isoform context of these excision events, particularly in the case of unannotated junctions. Future experiments with long-read sequencing will be needed to both assign the disease-associated splicing events to their effector isoforms, as well as to discern the impact of alternative 5’ and 3’ UTR excision on downstream molecular phenotypes such as translation efficiency and RNA stability. While our splicing genetic scores were derived from individuals of European ancestry, we utilized two Biobanks, including the UKBB to provide power to our analyses, and also chose to retain the comprehensive genetic diversity captured in the All of Us cohort, owing to its greater representation of diverse ancestries. Though high sharing of sTWAS associations between the cohorts underscores the robustness of our results, we expect our scores to translate less well across non-European genetic ancestries given that SNPs in LD with causal SNPS, rather than just those that are causal alone can be used by the Bayesian ridge model to compute the splicing scores. This was quantified through β-attenuation, which was mild for our high confidence fine-mapped results, but more severe when subsetting to all results from the European-ancestry subcohort analysis. Increased availability of RNA-seq from diverse individuals will help to produce genetic scores that are more generally applicable to a broader range of individuals, as well as prioritize causal variants and transcriptomic phenotypes.

In summary, our results highlight splicing as a pervasive and independent mediator of genetic risk, complementing gene expression in shaping the transcriptomic basis of complex traits. Through fine-mapping splicing-disease associations and by jointly integrating splicing with expression across two large biobank cohorts, we provide mechanistic resolutions for GWAS loci and demonstrate the utility of modelling multiple transcriptomic modalities in parallel. These insights underscore the importance of incorporating splicing into future large-scale functional genomics efforts, and suggest that splicing-aware approaches will be essential for advancing disease biology, therapeutic target discovery, and precision medicine.

## Data availability

The INTERVAL study data used in this paper are available to bona fide researchers from. The data access policy for the data has been approved by the ethics committee and is available at https://www.donorhealth-btru.nihr.ac.uk/research/blood-donors-bioresource/. This study used data from the All of Us Research Program’s Controlled Tier Dataset version 7, available to authorized users on the Researcher Workbench (https://researchallofus.org/). UK Biobank data are available (http://www.ukbiobank.ac.uk/) upon application and with permission of UKBB’s Research Ethics Committee. This research was conducted using the UKBB Resource under application no. 47976. Total gene expression scores were downloaded from the OmicsPred portal (https://www.omicspred.org/). ENCODE RNA binding protein eCLIP data was downloaded from the ENCODE portal (https://www.encodeproject.org/eclip/). Trained splicing prediction models, as well as full sTWAS summary statistics are publicly available on Zenodo (https://zenodo.org/records/17593872). The full fine-mapping summary statistics are also available on Zenodo (https://zenodo.org/records/17594375).

## Code availability

All original code from this analysis has been deposited to GitHub (https://github.com/atokolyi/sTWAS_paper_scripts).

## Acknowledgements

We gratefully acknowledge All of Us participants for their contributions, without whom this research would not have been possible. We also thank the National Institutes of Health’s All of Us Research Program for making available the participant data examined in this study. We would also like to thank the participants of the UKBB and INTERVAL cohorts for their valuable contributions. This research was conducted using the UKBB Resource under application no. 47976. This work uses data provided by patients and collected by the NHS as part of their care and support. Additionally, this work was made possible by support from the MacMillan Family and the MacMillan Center for the Study of the Non-Coding Cancer Genome at the New York Genome Center. We would also like to thank the New York Genome Center Systems Biology team for their computational cluster support and results portal hosting.

## Author contributions

A.T conducted the statistical analyses, created figures, and wrote the paper. A.T, D.A.K, and S.B conceived the study design. S.B assisted in the implementation of B-LORE. D.A.K supervised the project. All authors reviewed and approved the final version of the paper.

## Methods

### Splicing data generation (INTERVAL)

The INTERVAL study consists of approximately 50,000 healthy adult blood donors recruited through England’s National Health Service Blood and Transplant (NHSBT). Genotypes and splicing quantification from a subset of these individuals (n=4,732) was obtained from the INTERVAL consortium as previously described^4^. Briefly, 830,000 variants were assayed on the Affymetrix Axiom UK Biobank genotyping array, phased using SHAPEIT3, and imputed using a combined 1000 Genomes Phase 3-UK10K reference panel. Imputation was performed via the Sanger Imputation Server (https://imputation.sanger.ac.uk) resulting in 87,696,888 imputed variants and lifted over to reference build GRCh38. Whole blood RNA was sequenced with 75 bp paired-end sequencing reads (reverse stranded) on a NovaSeq 6000 system (S4 flow cell, Xp workflow; Illumina). Reads were aligned to the GRCh38 human reference genome (Ensembl GTF annotation v99) using STAR (v2.7.3a). The STAR index was built against the GRCh38 Ensembl GTF v99 using the option -jdbOverhang 75. STAR was run in a two-pass setup with standard ENCODE options to increase mapping accuracy and detect novel junctions. Splice junctions were extracted from aligned RNA-seq BAMs for the 4,732 individuals using Regtools (v0.5.2) junctions extract. Introns represented by extracted splice junctions were then clustered into groups using the Leafcutter^39^ pipeline (v0.2.9). Clustered introns were then prepared for sQTL analysis using “prepare_phenotype_table.py” to convert intron counts to normalized ratios. Total observed introns were filtered to those that were autosomal and overlapping an expressed gene body, with CPM > 0.5 in at least 24 individuals, and sufficient variance (minimum two filtered splicing event phenotypes per cluster), resulting in 111,937 filtered splicing event phenotypes in 11,016 genes. Splicing events were then annotated for transcriptomic and proteomic features using the Leafidex tool (https://github.com/atokolyi/Leafidex).

### Splicing genetic score training & evaluation

The full INTERVAL cohort was subset in to an 80% train subset (n=3,786) and a 20% withheld test subset (n=946), and sQTLs re-called with tensorQTL^40^ on the train subset in *cis* (+/-500kb from the center of the splice site, minor allele frequency (MAF>0.005, FDR<0.05) and *trans* (genome-wide, MAF>0.005, Bonferroni corrected). Both *cis* and *trans* analyses used previously defined covariates^4^, namely demographic variables, technical variables, and genotype and splicing PCs. For genetic score training *cis*-or *trans*-significant sQTLs were filtered to those that were regulating splice junctions outside the extended MHC region^17^ and were strictly unambiguous, bi-allelic single-nucleotide polymorphisms, resulting in 1,017,820 unique SNPs. To minimise redundant information and speed up computation, these SNPs were pruned with plink2^41^ “indep-pairwise” using a window size of 1000kb and an R^2^ threshold of 0.8, resulting in 234,889 pruned SNPs associated with 22,871 splice junctions for model training.

Splicing phenotypes for the 22,871 splice junctions were residualized using the above sQTL covariates. Bayesian Ridge (BR) regression was used in python (3.13.1) with sklearn^42^ (1.6.1) “BayesianRidge” to re-estimate sQTL effect betas, using uninformative priors (10e-5) as these have been previously demonstrated as optimal for the prediction of transcriptomic traits^10^. These residualized splicing genetic models were validated using 5-fold cross validation on the 80% subset (internal validation), with and without the addition of trans-sQTL SNPs to evaluate the added benefit of their inclusion. The combined *cis* & *trans* sQTL models were then applied to the 20% withheld subset to evaluate the trained models on unseen samples (withheld test subset validation). Of the 22,871 tested splice junctions, 13,915 had a mean withheld test set R^2^>0.01, and of these 13,851 were imputable in both the UKBB and AoU due to SNP overlap.

### UK Biobank data processing & imputation

To provide a comparable genetic background to that of INTERVAL, we restricted UKBB to the white-British (WB) subset, and individuals whose reported sex matched the genetically-inferred sex (n=408,590) to prevent ambiguity in sex-specific PheCode analysis, using the post-quality controlled genotype data^12^. Health PheCodes were extracted from the UKBB using the University of Michigan Center for Precision Health Data Science “createUKBphenome” pipeline (https://github.com/umich-cphds/createUKBphenome). PheCodes allow for the collapsing ICD codes into phenotypes of interest, to define individuals with a shared health phenotype, as well as provide related phenotypes to exclude from the control population for phenome-wide association testing^14,15^. We restricted the analysis to PheCodes with at least 200 cases in both the UKBB and AoU, resulting in 1,129 tested phenotypes. To impute the residualized splicing phenotypes into UKBB, we retrained the BR models for the 13,851 phenotypes passing withheld test set validation on the full INTERVAL cohort (n=4,732) using the subset of SNPs overlapping with UKBB (99.51%) and re-pruned as above. These trained models were then used for prediction with the UKBB genotypes, and the output predicted splicing phenotypes were residualized against the top 17 UKBB genotype PCs to minimize the effects of differing population structure on subsequent results.

### All of Us data processing & imputation

Although our models were trained in European-ancestry individuals, we applied them to the full All of Us v7 cohort^13^ to assess the broader generalizability of our results. The analysis was restricted to individuals who had both electronic health records (EHR) and short-read WGS data, and individuals whose reported sex matched the genetically-inferred sex (n=201,958). This included individuals of predicted genetic European (EUR n=114,249), African (AFR n=44,650), Admixed American (AMR n=35,564), East Asian (EAS n=4,178), South Asian (SAS n=2,529), and Middle Eastern (MID n=788) ancestry. PheCodes were mapped from ICD9-CM and ICD10-CM using a custom pipeline referencing the PheWAS catalog database^15^ as above (version 1.2), using the 1,129 qualifying PheCodes. Splicing phenotypes were imputed using BR models retrained on the full INTERVAL cohort using all SNPs (as AoU used short-read WGS, 100% of INTERVAL SNPs were retained) and re-pruned as above. Predicted splicing events were then residualized against the top 17 genotype PCs calculated from the short-read WGS data to minimize the effects of differing population structure on the results. As shown in **Figure S6**, the resulting p-values in the full cohort are generally lower (more significant) than when restricting to European-ancestry individuals, reflecting increased sample size combined with shared predictive signal across ancestries.

### Splicing transcriptome-wide association study

Next, a splicing TWAS was performed independently in UKBB and AoU due to their separate research analysis platforms by associating the 1,129 PheCodes with 13,851 predicted residualized splicing phenotypes which could be imputed in both cohorts, using a logistic regression model with statsmodels^43^ (0.14.4) in python (3.13.1), with sex and age as covariates, resulting in 16,496,541 tests. Analyses were restricted to be sex-specific when required, as denoted by the PheWAS catalog database. Test statistics and p-values were extracted, and p-values FDR (Benjamini & Hochberg) corrected.

An ancestry-stratified analysis was performed in the All of Us biobank to calculate the β correlations and attenuation ratios as a quantitative proxy for splicing model predictive accuracy (withheld R^2^) loss, to validate the European ancestry-trained models in non-European populations due to expected differences in LD structure. To perform this, we stratified individuals of genetic ancestry groups with sample size>10,000 (EUR n=114,249, AFR n=44,650, AMR n=35,564), and re-ran the logistic regression model for all qualifying PheCodes with sub-cohort case n≥200 as above, and extracted test statistics from these models. We quantified cross-ancestry attenuation of TWAS effect sizes by estimating a scaling parameter λ using inverse-variance weighted (IVW) regression of African-ancestry (AFR) effect sizes on European-ancestry (EUR) effect sizes within All of Us, using signals subsetted as ‘high-confidence’ (meta-analysis FDR<0.05 and PIP>0.5), or those FDR<0.05 in the AoU EUR sub-cohort.

A fixed-effects meta-analysis was then conducted in R (4.4.2) to combine the results from both cohorts, and the resulting p-values FDR (Benjamini & Hochberg) corrected. To test the robustness of our model R^2^>0.01 inclusion threshold, we performed a sensitivity analysis. We stratified all splicing models into four additional bins based on their validation R^2^ (>0.05, >0.10, >0.15, and >0.20). We then subsetted our sTWAS using only the splicing events in each bin and calculated the FDR (Benjamini & Hochberg) corrected (p<0.05) proportion of significant sTWAS associations (Number of Hits / Number of Models Tested) for each threshold.

### Joint Bayesian fine-mapping of sTWAS results with total gene expression

As TWAS effects may be in-part driven by linkage-disequilibrium and pleiotropy effects, we conducted a joint fine-mapping of splicing events by adapting a Bayesian multiple logistic regression framework (B-LORE)^16^. We opted to adapt B-LORE over existing tools such as FOCUS^25^ which rely on summary statistics, as our sTWAS approach uses the full matrices of predicted splicing and gene features, and PheCode case/control status, to avoid the unnecessary computation of full summary statistics for each PheCode. Additionally B-LORE outperforms the standard multiple linear regression models when nonlinearities become strong, such as in the case of imbalance case and control ratios in biobanks. Previously published total gene expression genetic scores^10^ (n=13,668) were used to predict gene expression features in the UKBB and AoU samples in the same methodology as for splicing events. Splicing and gene expression features were assigned to previously defined LD blocks^44^. A joint B-LORE model was run separately for each PheCode and cohort, for all significant PheCodes from the sTWAS meta-analysis, using all gene expression and splicing features contained in an LD-block together as a joint group, with age and sex as covariates and default parameters. There were 266/4966 splicing events for which the PheCode B-LORE model did not converge in either the UKBB or AoU, and hence were excluded from the meta-analysis. A B-LORE meta-analysis on the results from both the UKBB and AoU was then performed to compute posterior inclusion probabilities (PIPs) for each splicing-PheCode effect using zmax=10 (maximum number of causal features per LD block), nominating causal results as those with a meta PIP>0.5. The majority (79%) of LD block-PheCode pairs had less than 10 nominated causal transcriptomic features, providing an appropriate upper bound to balance excessive computation and prioritise stronger effects downstream.

---

**Figure S1.**
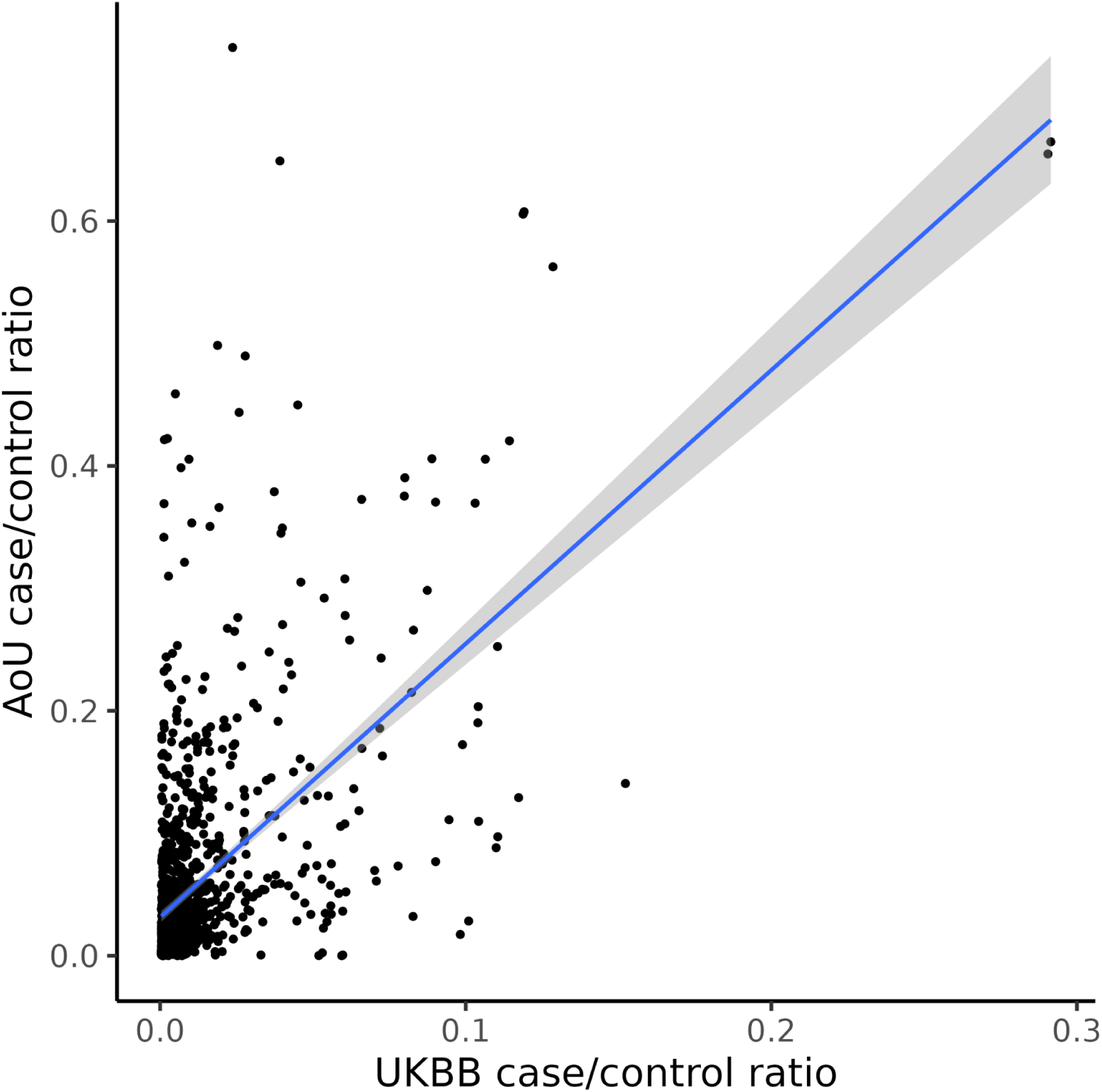
Comparison of case/control ratios between the UK Biobank and All of Us biobank. Points represent individual PheCodes, and the blue line represents a line of best fit, with a shaded 95% confidence interval.

**Figure S2.**
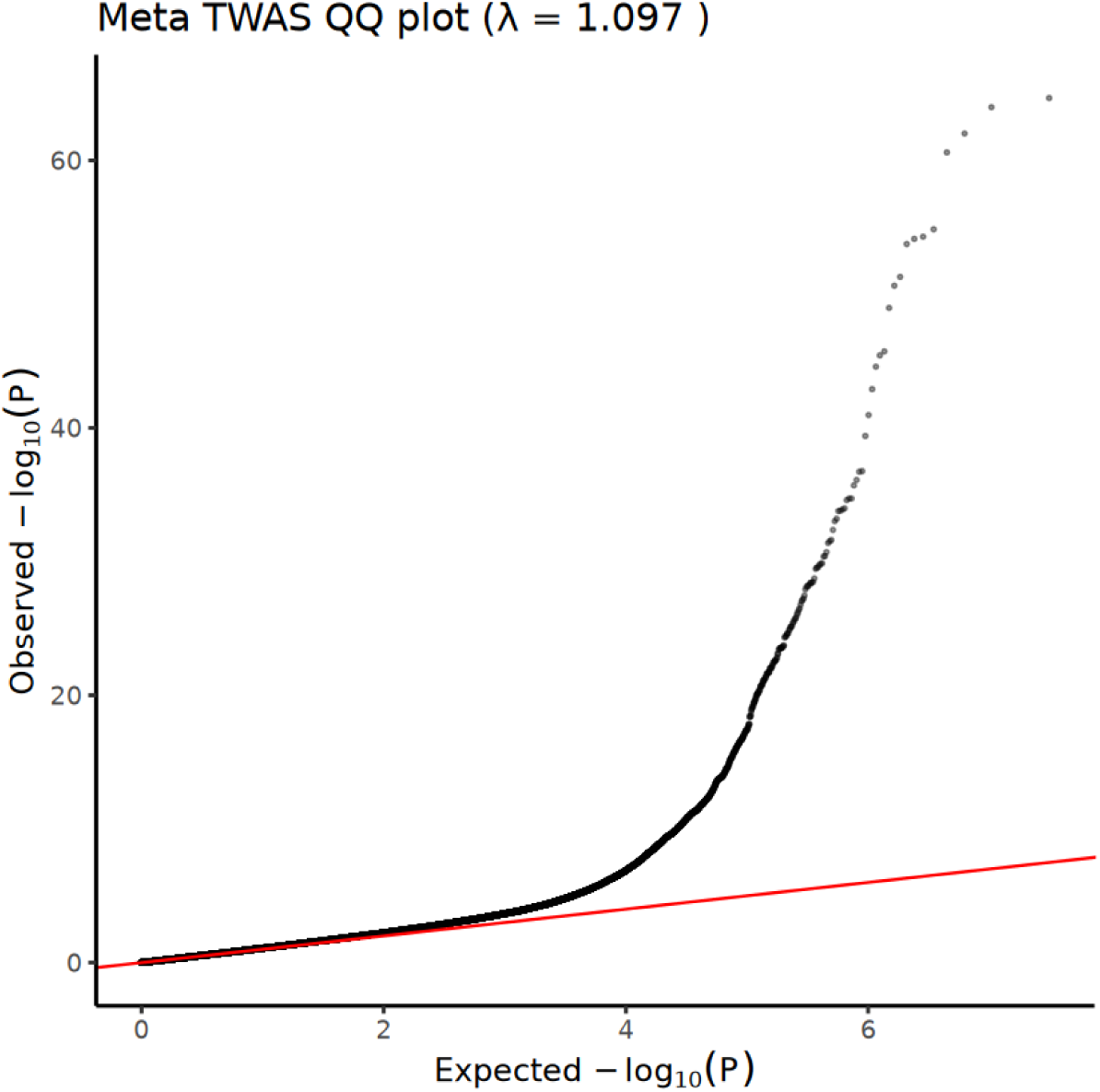
QQ plot of TWAS meta-analysis association statistics. Red line represents the expected null distribution, and black points represent the observed −log10(p) values.

**Figure S3.**
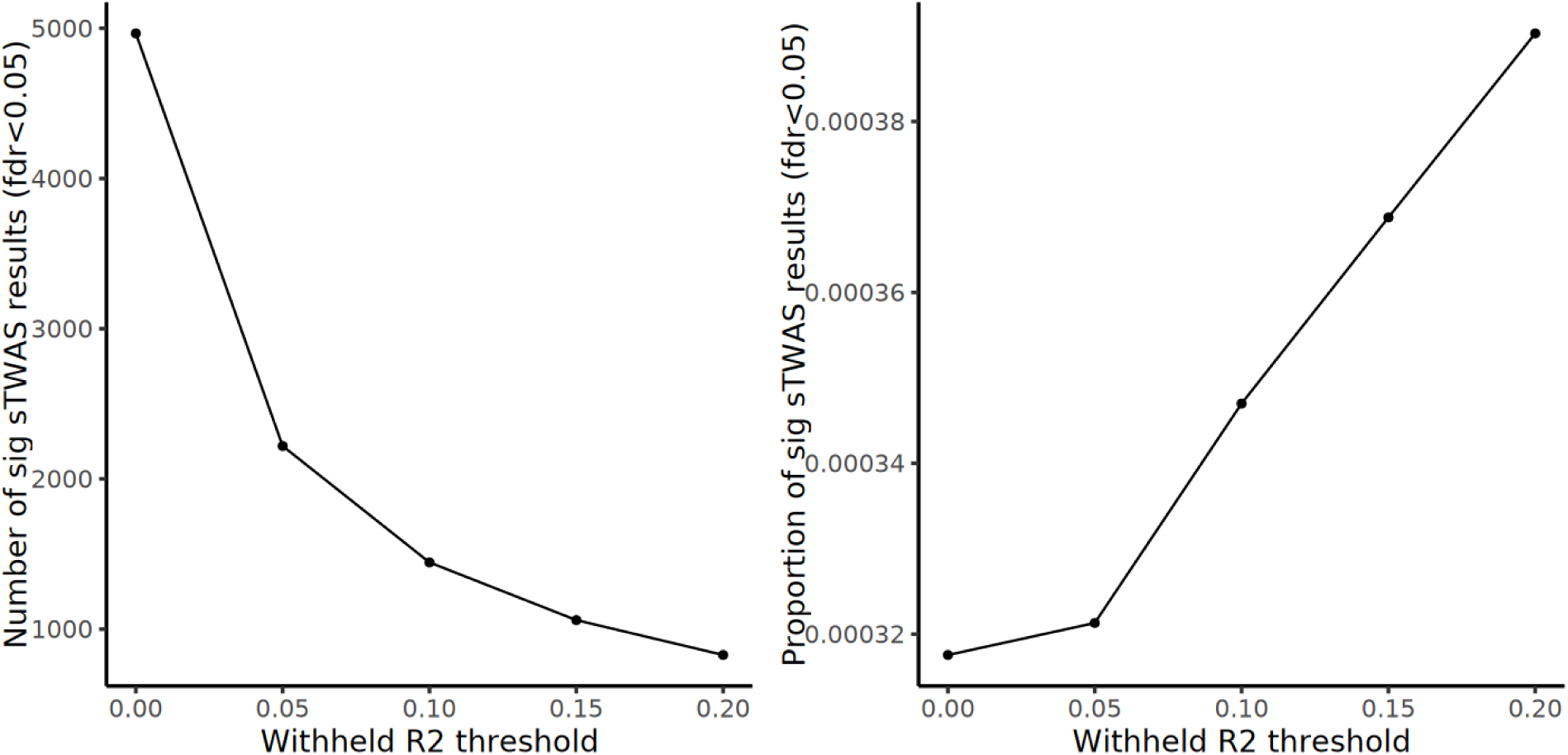
Sensitivity analysis for sTWAS with increasingly stringent R^2^ thresholds for withheld test-set accuracy for the inclusion of predicted splicing features.

**Figure S4.**
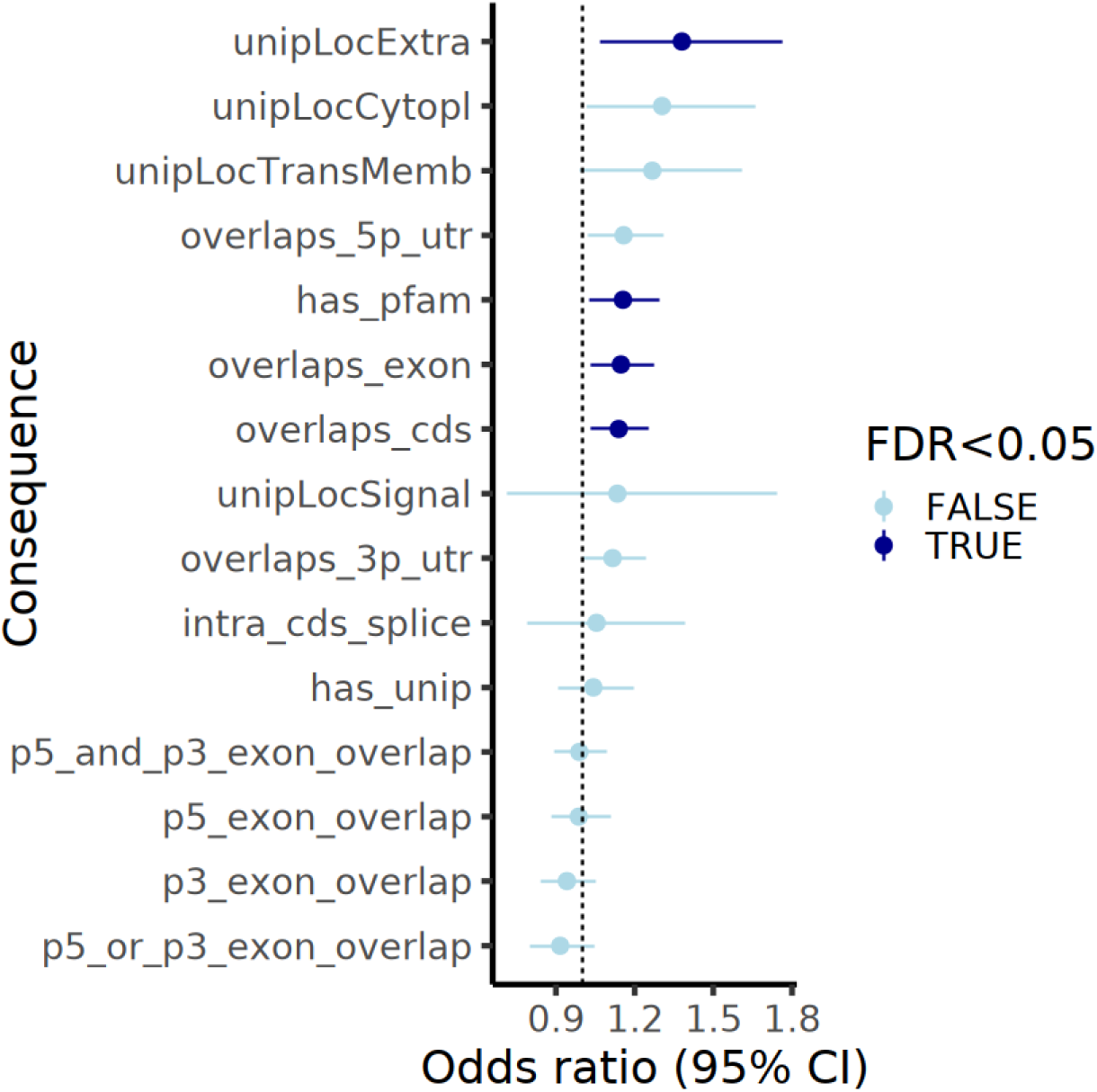
Enrichment of annotations for splicing events with a significant sTWAS association. Points represent odds ratios, with extending lines showing the 95% confidence interval. Those that are significant at FDR<0.05 are shaded darker.

**Figure S5.**
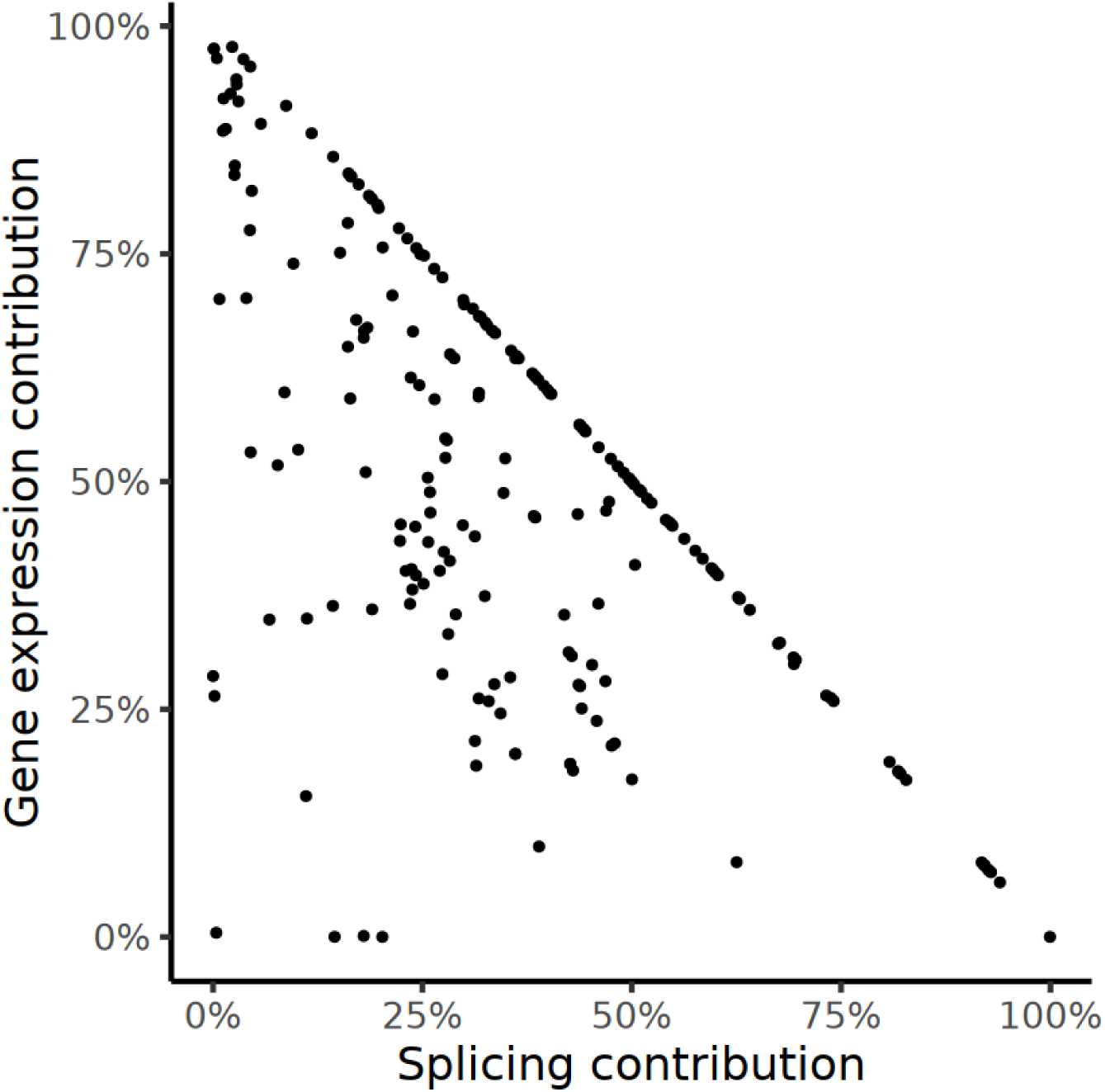
Scatter plot showing the relative transcriptomic proportion of the contribution of splicing and gene expression to traits. Each point represents a combined gene expression and splicing model of a single PheCode. Points below the y= -x line are those which have a shared component.

**Figure S6.**
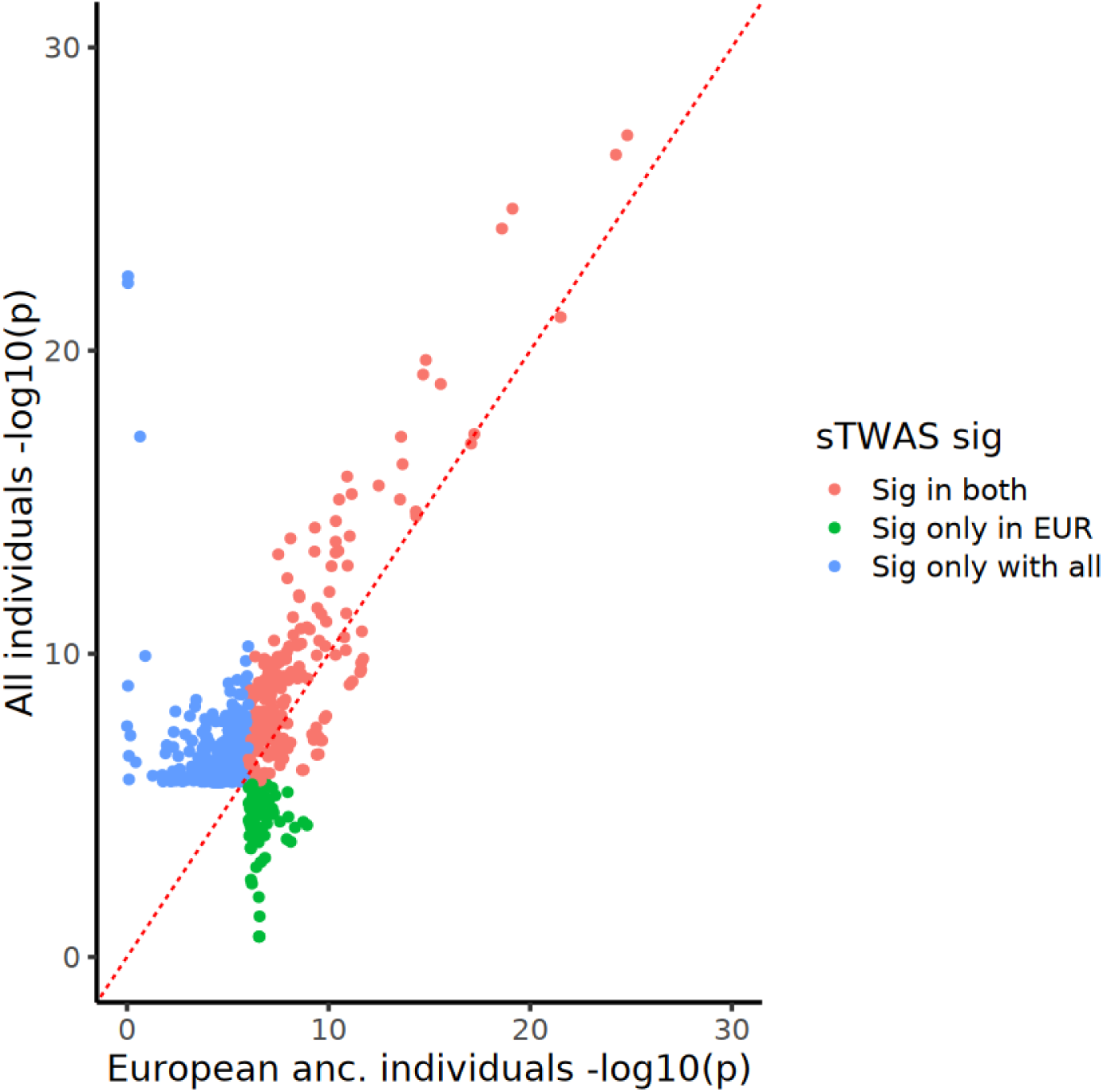
Comparison of AoU TWAS association significance between European-only and full All of Us cohort. Red dashed line represents y=x, splicing event-PheCode pairs which are significant in both analyses are in red, those only significant in the European-only subset are in green, and those only significant in the full AoU cohort are in blue. Pairs that are not significant in either analysis are omitted from the plot.

## Notes

### Competing Interest Statement

The authors have declared no competing interest.

### Author Declarations

North West Multi-Centre Research Ethics Committee of the NHS National Research Ethics Service gave ethical approval for UK Biobank (Ref: 11/NW/0382). All of Us Institutional Review Board of the National Institutes of Health gave ethical approval for the All of Us Research Program (Protocol ID: 2016-PM0001). National Research Ethics Service Committee East of England - Cambridge South gave ethical approval for the INTERVAL study (Ref: 11/EE/0538).

### Summary of Updates

Added new analyses exploring sensitivity, beta-attenuation between ancestry groups, and test statistic calibration. Minor revisions were made to the title, abstract, and discussion. This is the final version that was submitted to a journal for peer review.

